# Assessing the efficacy of Chlorella vulgaris for Vitamin B12 Deficiency: A Randomized Controlled Trial

**DOI:** 10.64898/2026.03.16.26348496

**Authors:** Subbu Kesavaraja, Sailaja Veluvali, Rajendran Lingan, Divya Chandradhara

**Author notes:** Corresponding Author: Name: Dr. Subbu Kesavaraja Address: EID Parry (Ind) Ltd, Parry Nutraceuticals, Dare House, 234, NSC Bose Road, Parry’s corner, Chennai, Tamil Nadu, India.

## Abstract

**Context:** Vitamin B12 deficiency is common among vegans and vegetarians due to limited intake of animal-derived foods. Identifying safe, plant-based sources of vitamin B12 is essential to address this nutritional gap.

**Aims:** This study evaluated the efficacy and safety of *Chlorella vulgaris* tablets in improving vitamin B12 deficiency.

**Settings and Design:** A double-blind, randomized, placebo-controlled trial was conducted among 46 healthy adults with vitamin B12 deficiency (serum levels 107–210 pmol/L).

**Methods and Material:** Participants were randomized (1:1) to receive *C. vulgaris* (1 g twice daily) or identical placebo for 12 weeks. Primary outcome was change in serum vitamin B12; secondary outcomes included folic acid, homocysteine, methylmalonic acid (MMA), and quality of life (WHO-QoL). Assessments were conducted at baseline and week 12, with safety monitored through liver and kidney function tests and adverse event reporting.

**Statistical Analysis Used:** Sample size (n=46) was calculated with 90% power and 10% dropout allowance. Data were analyzed using SPSS v22. Non-parametric tests were applied after normality assessment, with p<0.05 considered significant.

**Results:** Of 46 participants (mean age 35.5 ± 11.2 years; 69.6% female), mean serum vitamin B12 levels were significantly higher in the *C. vulgaris* group than in the placebo group at 12 weeks (214.4 ± 160.8 vs 55.9 ± 15.0 ng/mL; *P* < .001). No significant differences were observed in folic acid, homocysteine, MMA, or QoL scores between groups. No adverse events were reported.

**Conclusions:** Supplementation with *Chlorella vulgaris* significantly improved serum vitamin B12 levels, suggesting its potential as a safe, plant-based alternative for managing vitamin B12 deficiency.

**Key Messages:**

1. Plant-based Chlorella vulgaris improved vitamin B12 levels significantly
2. Randomized trial in B12-deficient healthy adults over 12 weeks
3. No adverse effects observed on liver or kidney function tests
4. Quality of life improved across all domains in the intervention group

## 1. Introduction

Vitamin B12 is an essential water-soluble vitamin involved in DNA synthesis, red blood cell formation, myelination, and neurological function. Deficiency, defined by low serum levels or impaired intracellular utilization, is highly prevalent in India, largely due to vegetarian dietary patterns and rising metabolic disorders. Plant-based alternatives are therefore increasingly sought. Microalgae, particularly *Chlorella vulgaris*, are rich in vitamin B12 and other bioactive nutrients. Despite its nutritional potential and sustainable cultivation advantages, clinical evidence supporting its efficacy as an oral B12 supplement remains limited. This study evaluates the effectiveness of *C. vulgaris* in improving serum vitamin B12 levels, related biomarkers, and quality of life.

## 2. Subjects and Methods

### 2.1 Study Design and Population

This study was a single-centric, randomized, double-blind placebo-controlled trial design that follows the CONSORT guidelines given by the EQUATOR network conducted at Sapthagiri Clintrac Private limited, Bengaluru, India. Participants with Vitamin B12 deficiency were enrolled from 13 February 2024 to 02 April 2024 with the trial assessments and follow up spanning 12 weeks. The research was conducted at Sapthagiri Clintrac Private limited with ethics approval obtained by the Sapthagiri Institute of Medical Sciences and Research Institutional Ethics Committee which was in accordance with the Declaration of Helsinki. Ethics approval number: SIMS&RC/IEC/AP-007/2023-24. The study was registered at the Clinical Trials Registry – India (CTRI) with the registration number CTRI/2023/08/056667. (weblink: ctri.nic.in/Clinicaltrials/pmaindet2.php?EncHid=OTEyODU=&Enc=&userName=). CONSORT reporting guidelines for randomised trial has been utilised ^1^.

### 2.2 Participants

46 participants were enrolled in the study based on the following criteria: Participants aged between 18–60 years; subjects who agreed to refrain from consuming dairy products; subjects with body mass index (BMI) between 18.5 and 30 kg/m²; subjects whose serum Vitamin B12 levels are between 107 and 210 pmol/L; subjects whose hemoglobin levels are ≥11 g/dL for women and ≥12 g/dL for men; females with a negative urine pregnancy test at the time of screening. Additionally, subjects with unstable medical conditions, chronic diseases (such as neurological, hepatic, renal, cardiovascular, gastrointestinal, pulmonary, or endocrine disorders), COVID-19 symtoms or those with malignancies that could confound study outcomes; subjects with a history of diabetes; subjects taking any other form of vitamin B12 supplements (either alone or in combination with other vitamins and minerals) or those who have received a vitamin B12 injection in the past 6 months were excluded from the study.

### 2.3 Randomization and Blinding

After obtaining written consent, participants were randomly assigned in 1:1 ratio to the study groups. Computer-generated sequence randomization was used. Allocation concealment was maintained using sequentially numbered, sealed, opaque envelopes provided to a nurse who was blinded to the group assignments. The nurse, otherwise not involved in the trial, was assigned the intervention according to the allocation sequence contained in each envelope. Participants, research team and data analysts were blinded to the treatment group.

### 2.4 Intervention

After randomization, the participants received placebo or intervention as per the allocation. The intervention group received 1 gram of proprietary dietary supplements, Parry’s Chlorella tablet twice daily before meals with water. Each gram of tablet contains 472.5μg per 100g of *C. vulgaris* dry powder along with several other bioactive compounds as detailed in the supplementary file below. The tablet contained whole-cell *C. vulgaris*, a green microalga rich in biologically active vitamin B12. While there are several methods for *C. vulgaris* production, in the present study algae were cultivated in naturally open raceway pond using organic nutrients without employing traditional cell wall cracking methods commonly used worldwide. This unique and proprietary cultivation method conducted in open race way ponds with high radiation conditions provides a favorable environment for the algae to accumulate biomass and enhance both cell density and nutrient composition of the cells ^2^. These tablets were manufactured at Parry Nutraceuticals with FSSAI Number: 10013042001015. The tablets did not contain any added sugars, starch, preservatives, or artificial colors. The placebo group received organoleptically similar tablets containing maltodextrin and chlorophyll. Both groups received their respective intervention for a period of 12 weeks (95 days). All the participants received a calculated quantity of intervention tablets during each visit. The accountability of consumed versus unused tablets was documented at each visit. Also, any instance of accidental or intentional drug destruction was recorded by the research staff based on participant reports. All unused tablets and other drug supplies were returned to the sponsor at the end of the study.

### 2.5 Data Collection

Baseline characteristics and clinical assessments such as physical examination, anthropometric measures, vital parameters, complete blood count, liver and kidney profile with other study-specific biochemical parameters were obtained from participants’ medical records. Changes in serum vitamin B== concentrations and other study-specific biochemical parameters were assessed at baseline and at the end of the 12-week intervention period (90 ± 2 days). Additionally, participants completed the World Health Organization Quality of Life (WHOQOL) questionnaire at baseline and at the end of the study. Safety evaluations were conducted at weeks 4 and 8 which included physical examination, vital sign measurements, documentation of adverse events, and evaluation of liver and kidney function parameters.

### 2.6 Study Outcomes

Study outcomes were assessed at baseline and after 12 weeks. The primary outcome was the statistically significant change in serum vitamin B12 levels from baseline to week 12 between the intervention and placebo groups.Secondary outcomes included any mean changes in blood biomarkers such as homocysteine, folic acid, and methylmalonic acid (MMA). Other secondary outcomes included improvement in Quality of life using the WHO Quality of Life (WHOQoL) BREF questionnaire. The scale is validated 5-likert scale for questionnaire with sub domains for physical health, psychological health, social relationships and environment ^3^.

### 2.7 Sample Size and Statistical Analysis

According to a prior study ^4^, assuming a study power of 90%, a standard deviation of 90, and a 10% dropout rate, a study sample size of 46 participants was sought. Statistical analysis was conducted using Statistical Package for Social Sciences software (SPSS) (version 22.0). Categorical variables were presented as counts and percentages and the continuous variables were presented as means with standard deviations or medians with interquartile ranges. Normality of the data was tested using the Kolmogorov Smirnov test. For parametric tests, the Wilcoxon signed-rank test was used to assess changes between the two groups, while the Mann-Whitney test was used to evaluate the significance between the time points. Statistical significance was defined as a p-value < 0.05.

Additionally, it evaluates the effect of Parry’s Chlorella supplementation on various biomarkers such as homocysteine levels, folic acid, methylmalonic acid (MMA), liver enzymes, lipid profile, and quality of life. The study spanned a 12-week duration which included a baseline visit followed by vital assessments at weeks 4, 8, and 12. The primary and secondary outcomes were assessed at baseline and week 12.

## 3. Results

Among the 56 participants identified for vitamin B12 deficiency, 46 participants (82.1%) were enrolled and randomized: 23 participants in the intervention group and 23 participants in the placebo group. Also, there were no dropouts in the study. A CONSORT flow diagram of the study protocol is represented in **Figure 1**. None of the participants reported any adverse events due to the intervention.

**Fig 1:**
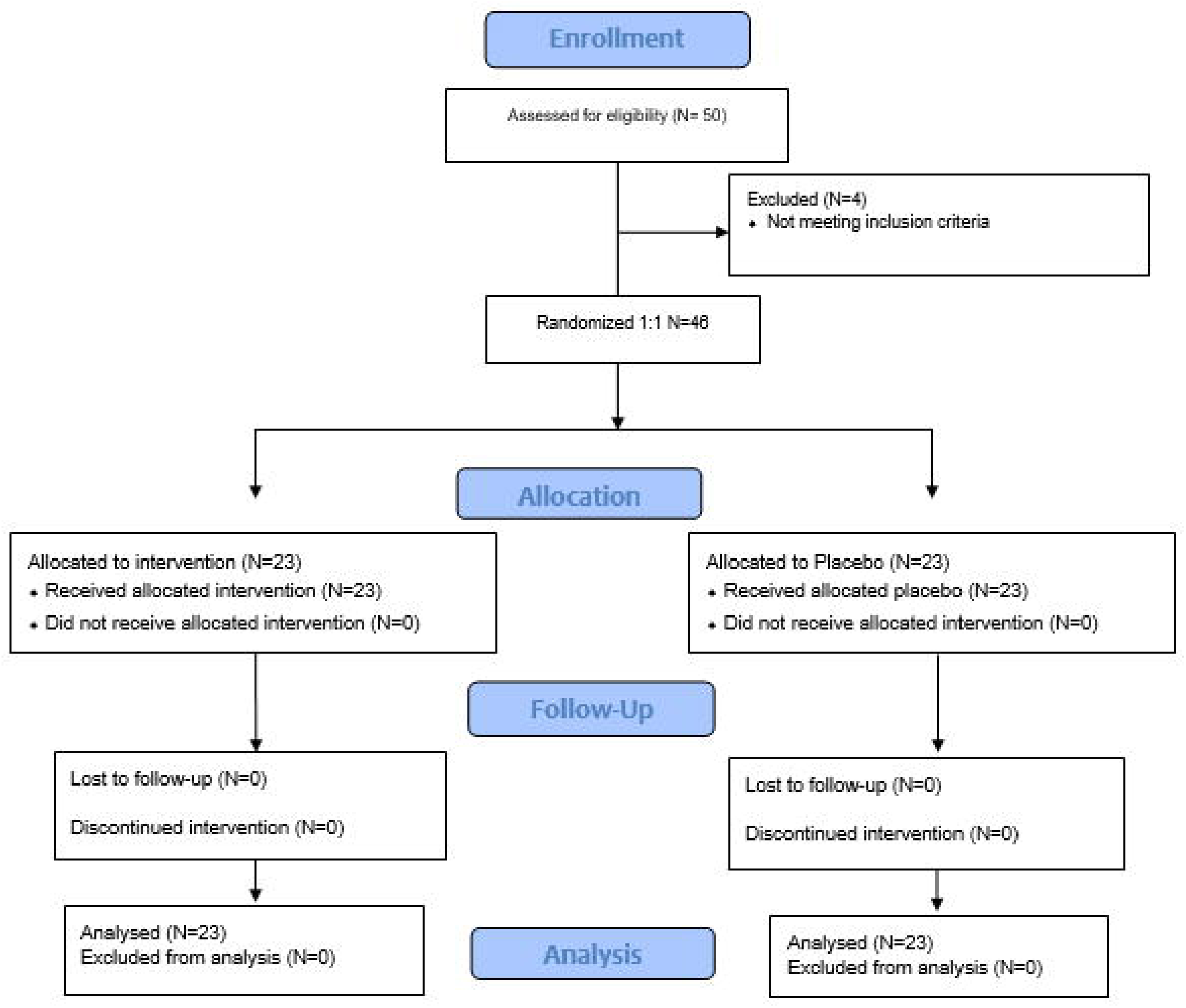
CONSORT Flow Chart of the randomization between Intervention and Placebo groups

### 3.1 Demographic and Baseline Characteristics

At baseline, the mean (SD) serum vitamin B12 concentration was 48.04 (7.96) ng/ml in the intervention group and 48.09 (9.56) ng/ml in the placebo group. The mean (SD) age was 35.52 (11.24) years in the intervention group and 35.57 (10.31) years in the placebo group. The male-to-female distribution was 30:60 in both groups, with a higher proportion of female participants overall. Other blood biomarkers such as baseline mean (SD) serum folic acid concentrations were 6.89 (3.98) ng/ml in the intervention group and 6.17 (2.94) ng/ml in the placebo group. Mean (SD) hemoglobin levels were 13.57(1.56) g/dL and 13.26 (1.63) g/dL in the intervention and placebo group respectively. Methylmalonic acid concentrations were 0.02 (0.01) (μmol/L) and 0.02 (0.02) (μmol/L), homocysteine levels were 21.2 (14.12) (μmol/L) and 18.72 (9.43) (μmol/L) in the intervention and placebo groups respectively. Both the groups did not show any significant differences in the baseline characteristics or anthropometric measurements as shown in Table 1.

**Table 1.** Baseline Demographic and Clinical Characteristics of Participants.

| Characteristic | Intervention Group<br>(n = 23) | Placebo Group<br>(n = 23) | *P* Value<sup>1</sup> |
| --- | --- | --- | --- |
| Age, mean (SD), y | 35.52 (11.24) | 35.57 (10.31) | 0.988 |
| Sex, No. (%) |  |  | 1.000 |
| Male | 7 (30.4) | 7 (30.4) |  |
| Female | 16 (69.6) | 16 (69.6) |  |
| Height, mean (SD), cm | 163.6 (6.53) | 161.8 (4.89) | 0.296 |
| Weight, mean (SD), kg | 64.35 (7.23) | 62.52 (8.48) | 0.435 |
| Body mass index, mean (SD), kg/m<sup>2</sup> | 24.08 (2.42) | 23.74 (2.79) | 0.661 |
| Systolic blood pressure, mean (SD), mm Hg | 119.6 (2.09) | 118.7 (3.44) | 0.291 |
| Diastolic blood pressure, mean (SD), mm Hg | 80 (3.02) | 78.7 (3.44) | 0.180 |
| Pulse rate, mean (SD), beats/min | 84.57 (1.73) | 85.39 (2.13) | 0.159 |
| Haemoglobin, mean (SD), g/dL | 13.57 (1.56) | 13.26 (1.63) | 0.513 |
| Haematocrit, mean (SD), % | 39.53 (4.96) | 38.43 (5.61) | 0.485 |

| Characteristic | Intervention Group<br>(n = 23) | Placebo Group<br>(n = 23) | P Value<sup>1</sup> |
| --- | --- | --- | --- |
| Vitamin B<sub>12</sub>,<br>mean (SD), ng/mL | 48.04 (7.96) | 48.09 (9.56) | 0.985 |
| Folate,<br>mean (SD), ng/mL | 6.89 (3.98) | 6.17 (2.94) | 0.489 |
| Methylmalonic acid,<br>mean (SD), μmol/L | 0.02 (0.01) | 0.02 (0.02) | 1.000 |
| Homocysteine,<br>mean (SD), μmol/L | 21.2 (14.12) | 18.7 (9.43) | 0.484 |

SD: Standard Deviation
p-value: p-value at baseline is comparison of actual values between groups
Continuous variables (such as age, height, weight, body mass index, blood pressure, pulse rate, hemoglobin, hematocrit, and biochemical parameters) were analyzed using the Welch's two-sample t-test, which does not assume equal variances between groups. This method was chosen because it provides a more robust estimate of group differences when standard deviations differ slightly or when sample sizes are relatively small but comparable (n = 23 per group).
For categorical variables (such as sex), intergroup comparisons were made using the Chi-square test of independence

### 3.2 Primary Outcome

At 12 weeks, mean (SD) serum vitamin B 12 concentrations were 214.4 (160.8) ng/mL in the intervention group and 55.9 (15.01) ng/mL in the placebo group (Table 2). The adjusted between-group difference was statistically significant (P<0.001, Mann Whitney test). From baseline to week 12, the mean (SD) change in vitamin B 12 concentration was 166.4 (159.7) ng/mL in the intervention group and 7.8 (16.7) ng/mL in the placebo group.

**Table 2.**
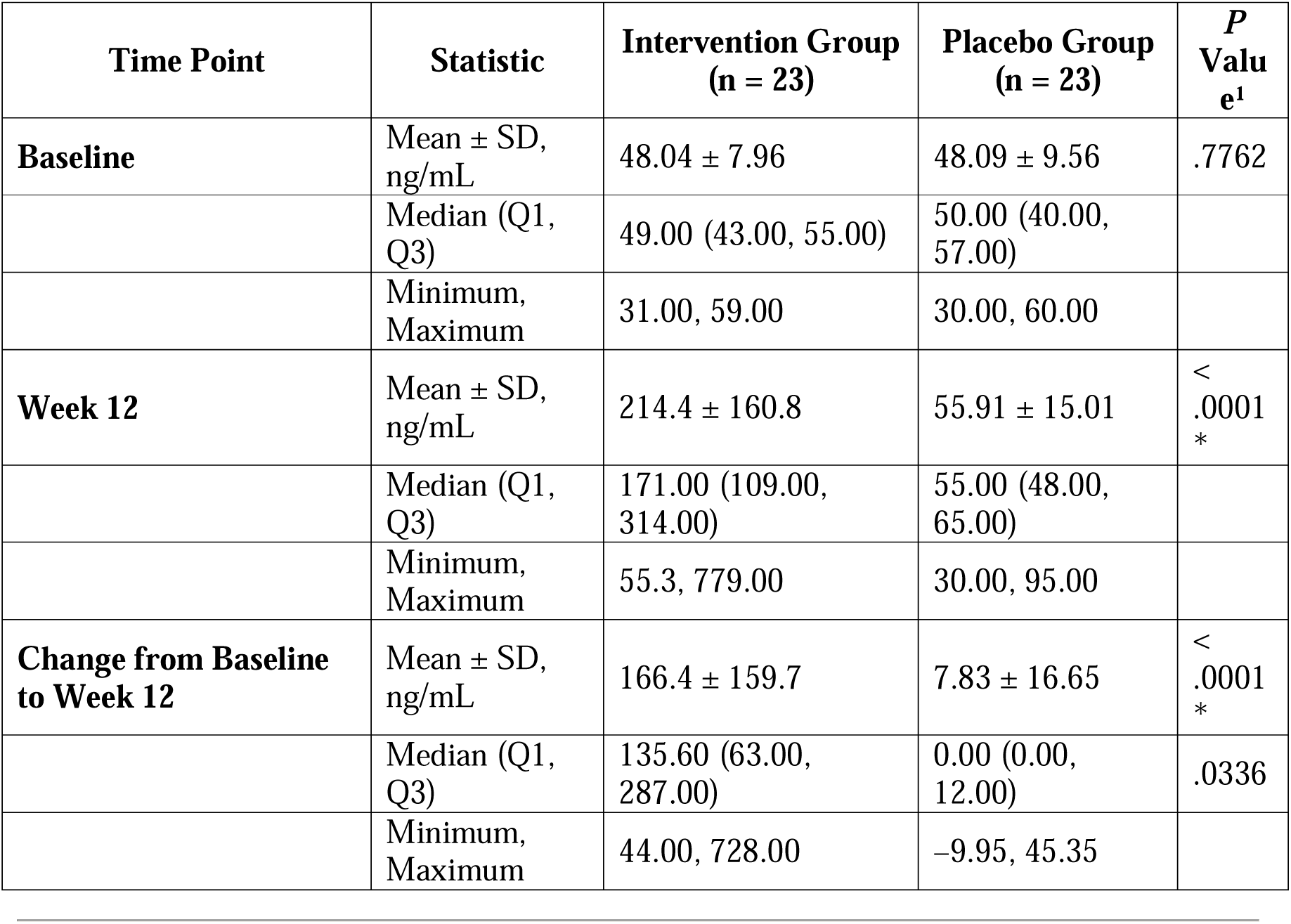
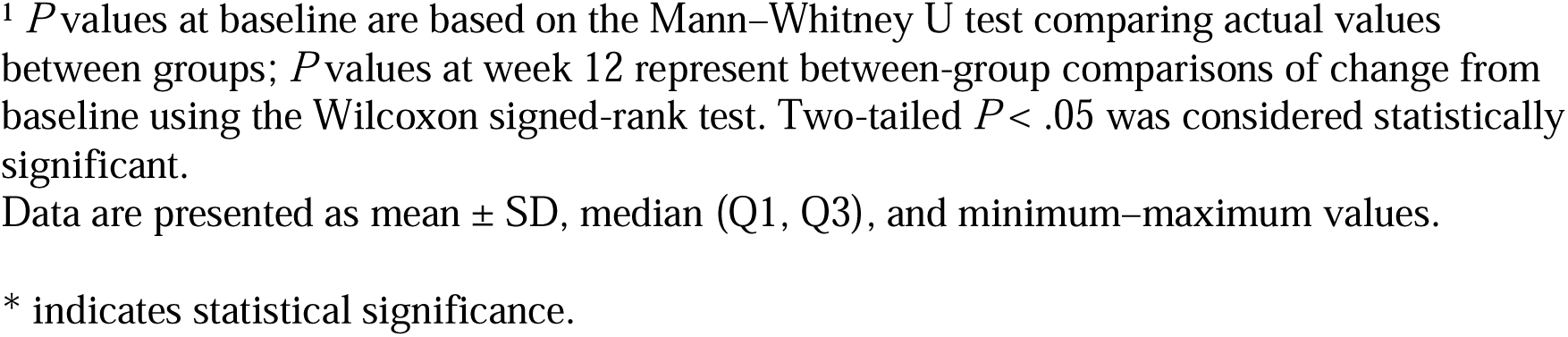
Serum Vitamin B. DD **Concentrations at Baseline and Week 12**

| Time Point | Statistic | Intervention Group<br>(n = 23) | Placebo Group<br>(n = 23) | P Value<sup>1</sup> |
| --- | --- | --- | --- | --- |
| **Baseline** | Mean ± SD, ng/mL | 48.04 ± 7.96 | 48.09 ± 9.56 | .7762 |
|  | Median (Q1, Q3) | 49.00 (43.00, 55.00) | 50.00 (40.00, 57.00) |  |
|  | Minimum, Maximum | 31.00, 59.00 | 30.00, 60.00 |  |
| **Week 12** | Mean ± SD, ng/mL | 214.4 ± 160.8 | 55.91 ± 15.01 | < .0001 * |
|  | Median (Q1, Q3) | 171.00 (109.00, 314.00) | 55.00 (48.00, 65.00) |  |
|  | Minimum, Maximum | 55.3, 779.00 | 30.00, 95.00 |  |
| **Change from Baseline to Week 12** | Mean ± SD, ng/mL | 166.4 ± 159.7 | 7.83 ± 16.65 | < .0001 * |
|  | Median (Q1, Q3) | 135.60 (63.00, 287.00) | 0.00 (0.00, 12.00) | .0336 |
|  | Minimum, Maximum | 44.00, 728.00 | -9.95, 45.35 |  |

<sup>1</sup> *P* values at baseline are based on the Mann–Whitney U test comparing actual values between groups; *P* values at week 12 represent between-group comparisons of change from baseline using the Wilcoxon signed-rank test. Two-tailed *P* < .05 was considered statistically significant.
Data are presented as mean ± SD, median (Q1, Q3), and minimum–maximum values.
\* indicates statistical significance.

### 3.3 Secondary Outcomes

Mean (SD) serum folic acid concentrations were 6.15 (4.07) ng/mL in the intervention group and 6.35 (2.14) ng/mL in the placebo group at week 12 (Table 3). The mean (SD) change from baseline was −0.74 (4.23) ng/mL and 0.18 (2.57) ng/mL, respectively. Between-group and within-group comparisons from baseline to week 12 showed no statistically significant differences (P >0.05, Wilcoxon signed-rank and Mann-Whitney tests). At 12 weeks, mean (SD) serum homocysteine concentrations were 19.34 (8.8) µmol/L in the intervention group and 23.68 (22.22) µmol/L in the placebo group. The mean (SD) change from baseline was −1.86 (16.64) µmol/L and 4.96 (21.84) µmol/L in the intervention and placebo groups, respectively, with no statistically significant differences between groups (P >0.05) (Table 3). At week 12, mean (SD) methylmalonic acid concentrations were 0.001 (0.01) µmol/L in the intervention group and 0.02 (0.04) µmol/L in the placebo group, with no meaningful change from baseline or significant between-group difference (P >0.05). While no significant changes were noted in secondary biochemical parameters, numerical reductions were noted in homocysteine levels and serum folic acid. MMA levels remained the same after the 12-week intervention. WHOQOL-BREF

**Table 3.**
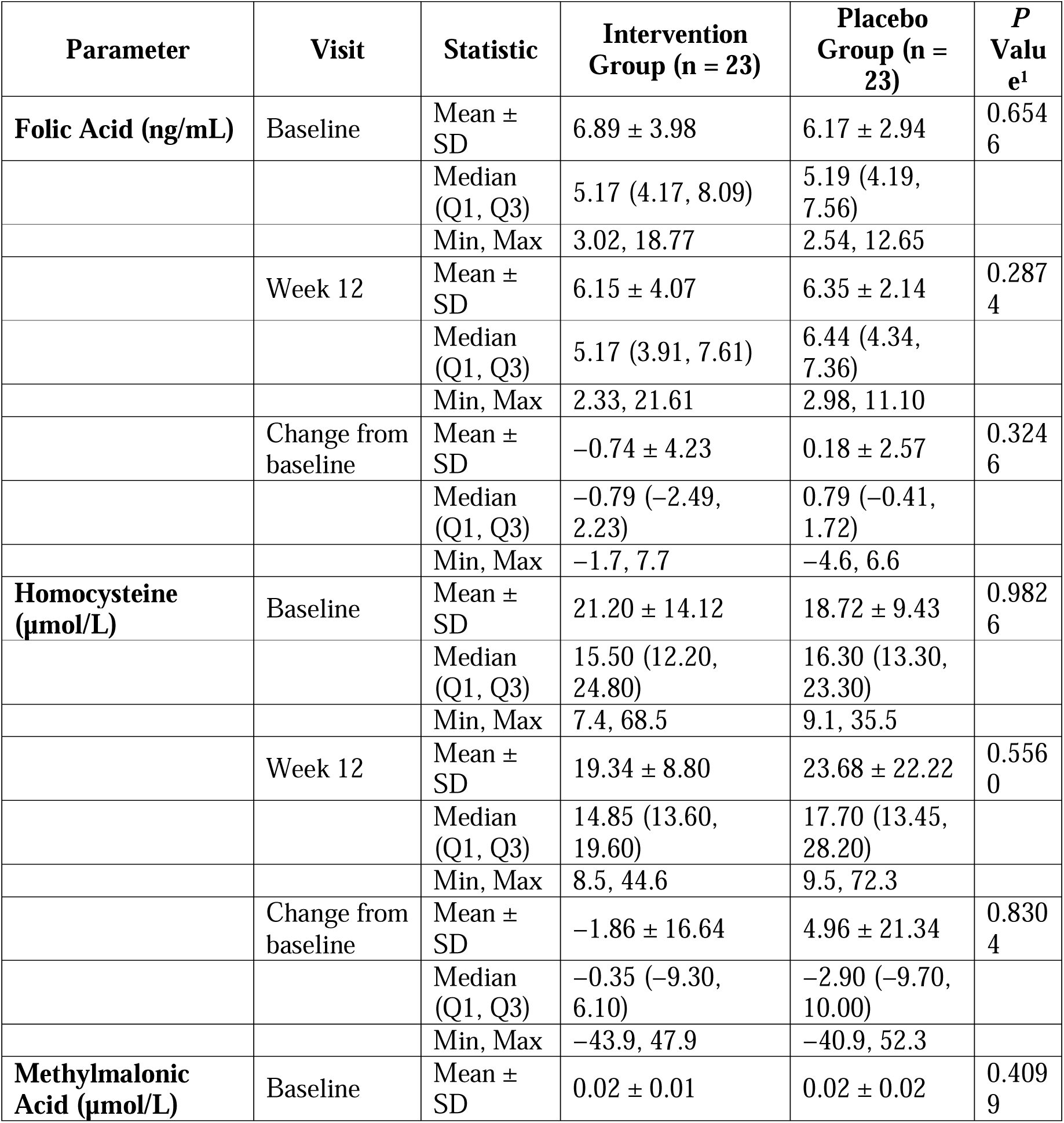
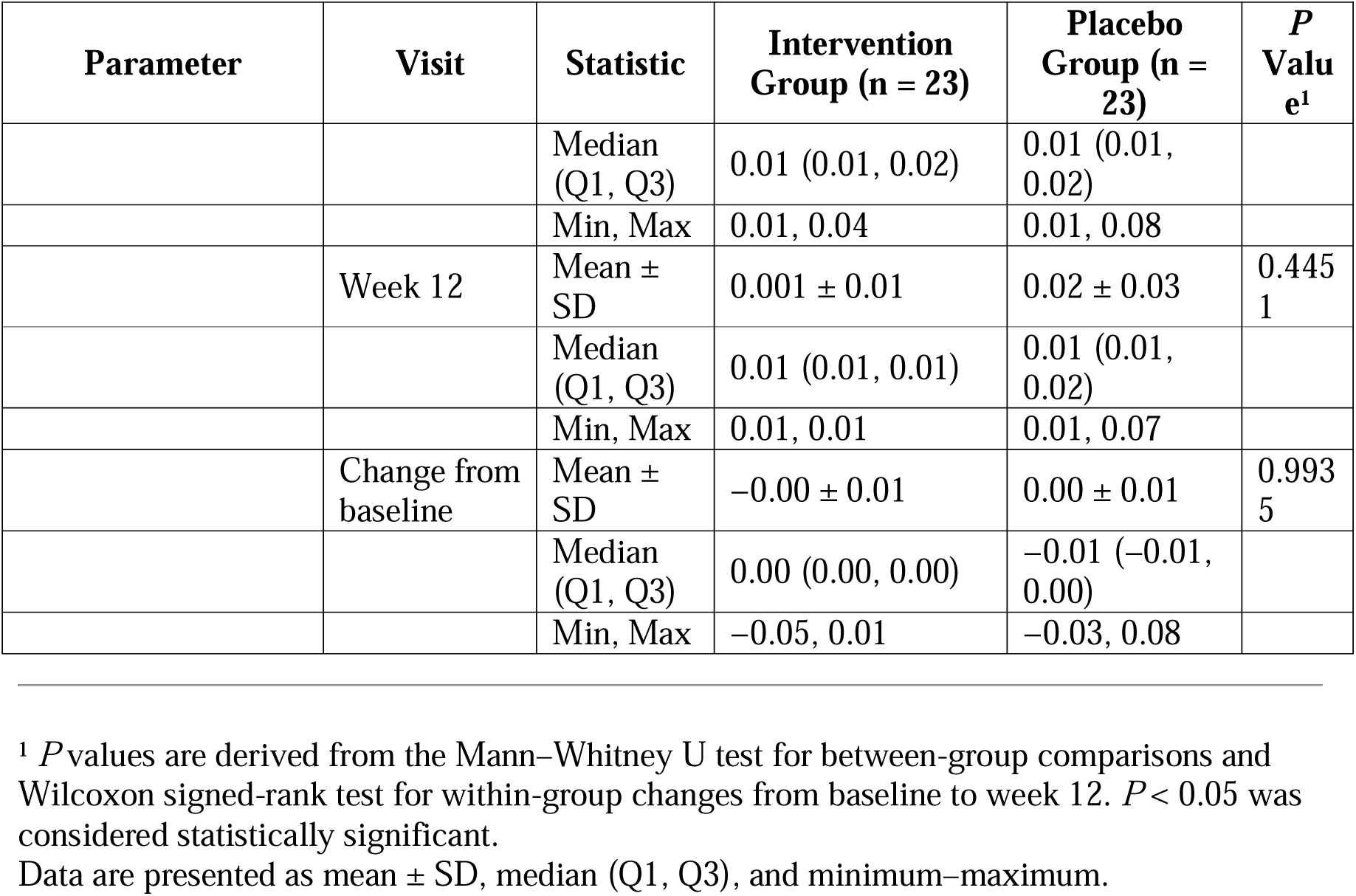
Summary of Serum Folic Acid, Homocysteine, and Methylmalonic Acid (MMA) Levels at Baseline and Week 12.

At 12 weeks, results from the WHOQOL-BREF questionnaire showed statistically significant improvements from baseline within both the intervention and placebo groups across several domains (Table 4). The mean (SD) change in physical health scores from baseline to week 12 was 0.61 (0.77) in the intervention group and 0.52 (0.77) in the placebo group (P =0.03 and P = 0.02, respectively). Psychological domain scores increased by 0.41 (1.08) in the intervention group and 0.44 (0.98) in the placebo group, both reaching statistical significance (P = 0.03 and P =0.02). The mean (SD) change in social relationship scores was 0.53 (1.13) in the intervention group and 0.52 (1.31) in the placebo group (P =0.048 and P=0.098). Only the intervention group reached statistical significance whereas the placebo group did not reach any statistical significance. Environmental domain scores improved by 1.59 (1.64) and 1.30 (1.18) in the intervention and placebo groups, respectively (P=0.001 and P<0.001). Although both groups demonstrated significant improvements from baseline across multiple domains, no statistically significant between-group differences were observed at 12 weeks for any of the WHOQOL domains (P >0.05).

**Table 4.** Summary of WHOQOL-BREF domain scores – change from baseline to week 12.

| Domain | Statistic | Intervention Group (n = 23) | Placebo Group (n = 23) | *P* Value<sup>1</sup> |
| --- | --- | --- | --- | --- |
| **Physical Health** | Mean ± SD | 0.61 ± 0.77 | 0.52 ± 0.77 | 0.0007*/0.0064* |
| **Psychological Health** | Mean ± SD | 0.41 ± 1.08 | 0.44 ± 0.98 | 0.03* / 0.02* |
| **Social Relationships** | Mean ± SD | 0.53 ± 1.13 | 0.52 ± 1.31 | 0.048 / 0.098 |
| **Environmental Domain** | Mean ± SD | 1.59 ± 1.64 | 1.30 ± 1.18 | 0.001* / <0.001* |

*p* values are derived using the Wilcoxon signed-rank test for within-group changes from baseline to week 12. Two-tailed *P* < .05 was considered statistically significant. Data are presented as mean ± SD unless otherwise indicated.

## 4. Discussion

The present study evaluated the efficacy and safety of Parry’s Chlorella tablets to improve the serum vitamin B12 levels among vegetarians. The study tested a total of 46 participants, 23 in each group randomly assigned to the intervention or placebo group. The study did not have any dropouts during the 12-week period and none of the participants reported any adverse events. The findings of the present study demonstrated a significant improvement in the vitamin B12 levels of the individuals in the intervention group as compared to those in the placebo group. Several studies in the past have highlighted the nutritional value of microalgae, especially Arthrospira and Chlorella species ^5^ supporting the findings of the present study. This increase in the vitamin B12 content was particularly noted in Chlorella as compared to Arthrospira because Chlorella species produces physiologically active form of vitamin B-12 (methylcobalamin) as compared to the pseudo-vitamin B12 ^6^. Another study quantified the methylcobalamin (Me-Cbl) content in *C. vulgaris* using microbiological and chemiluminescence methods. The study found that the Me-Cbl content was 29.87 ± 2 μg/100 g using the microbiological method and 26.84 ± 2 μg/100g using the chemiluminescence method. These findings indicate that *C. vulgaris* was a potent source of vitamin B12 and could be utilized as a nutritional supplement ^7^.

In the present study, the secondary outcome parameters such as homocysteine, folic acid and MMA levels, did not demonstrate significant differences between the intervention and placebo groups. Although serum vitamin B12 levels is commonly regarded as a biomarker for vitamin B12 deficiency, a more sensitive method of deficiency is measurement of homocysteine levels and MMA levels in the body. This is due to the fact that functional vitamin B12 are essential for conversion of methylmalonic acid (MMA) to succinyl-CoA, and conversion of homocysteine (Hcy) to methionine in the presence of vitamin B12 and folic acid ^8^.

However, homocysteine levels decreased in the intervention group, whereas in the placebo group an increase in homocysteine levels were noted. Since vitamin B12 plays a crucial role in the metabolism and breakdown of homocysteine, reduction in homocysteine levels in the intervention group could be attributed to a rise in vitamin B12 levels due to the intervention ^9^. Previous studies have documented the association of elevated levels of homocysteine with metabolic disturbances, including cardiovascular disease, stroke, and obesity which demonstrates the clinical relevance of vitamin B12 supplementation in managing homocysteine-related health risks ^9,10^. Folic acid, a precursor to vitamin B9, is closely interrelated with vitamin B12 levels ^11^. In the present study, folic acid levels slightly decreased in the intervention group, while they increased in the placebo group. The observed reduction in folic acid levels in the intervention group may be due to enhanced utilization of folic acid by increased levels of vitamin B12. The increase in folic acid levels in the placebo group could be attributed to a deficiency in vitamin B12, which impairs folic acid metabolism. Studies also interlink excessive folic acid in the body can increase vitamin B12 deficiency, potentially leading to other adverse clinical outcomes ^12^. These findings highlight the complex interplay between vitamin B12 and folic acid metabolism and the need for careful management of both nutrients. Elevated levels of Methylmalonic acid (MMA) is commonly observed in individuals with vitamin B12 deficiency ^13^. Increased MMA levels contribute to cardiovascular mortality risk, particularly in patients with coronary artery disease and liver disease ^13,14^. The present study observed comparable baseline MMA levels across both groups. After 12 weeks of intervention, MMA levels decreased in the intervention group, while the placebo group showed no change. Although the difference in MMA levels between the two groups was not statistically significant, even a modest reduction of 0.1 umol/L in MMA levels can be impactful on the well-being of individuals ^15^. Vitamin B12 is essential for numerous physiological functions, and its deficiency can significantly reduce the quality of life among individuals with deficiency. Studies have identified a range of physical and mental symptoms associated with vitamin B12 deficiency that detrimentally affect an individual’s well-being ^16^. These include mood swings, chronic fatigue, attention deficit disorder, as well as cognitive and behavioral changes ^17^. Previous studies have also demonstrated that addressing vitamin B12 deficiency may lead to improvements in conditions such as fibromyalgia, with notable reductions in constant pain, highlighting the potential benefits of vitamin B12 supplementation in managing this condition ^18^. The present study demonstrated that participants in the intervention group showed significant improvements in physical, psychological, social and environmental well-being, as assessed by the WHO Quality of Life (QOL) questionnaire, after 12 weeks. However, the improvement in self-reported questionnaires were also noted for both the groups indicating nonspecific or placebo-related responses rather than the intervention itself. The observed improvements in physical and psychological health following Chlorella consumption for the intervention group are consistent with previous research demonstrating improvement in fatigue and depression ^19^. Further, physical and mental health might have a beneficial effect on the social and environmental well-being ^20^ as noted in this study.

## 5. Strengths and Limitations

The strength of this study lies in its robust design; however, the sample size was relatively small, due to which significant changes in many of the secondary markers were not noted. Future studies with larger sample sizes could provide more definitive insights into the efficacy of *C. vulgaris* in several biochemical functions, further demonstrating its true potential for public health benefits. Another limitation of the study was that the dose-dependent effects of the dietary supplement were not studied, particularly in terms of its impact on vitamin B12 levels. Further randomized controlled longitudinal studies can assess the sustained impact of *C. vulgaris* supplementation. This may also help in understanding the optimal dosing regimen for improving vitamin B12 levels over time. Another limitation of the study lies in the cohort selected which included only mild patients with vitamin B12 deficiency. Further studies with a diverse cohort and varying deficiency levels can offer broader applicability. Additionally, evaluating the efficacy of the Chlorella dietary supplementation among pregnant women with vitamin B12 deficiency can be more impactful with reducing the risk of neurological disabilities in their offsprings or stillbirth.

## 6. Conclusion

Vitamin B12 deficiency, is a prevalent concern worldwide, particularly among individuals following plant-based diets. The present study highlights Chlorella, a microalga rich in micronutrients, as a promising plant-based candidate for addressing vitamin B12 deficiency. The study highlights the increase in vitamin B12 along with associated favorable outcomes and quality of life in the intervention group after the consumption of Chlorella tablets for 12 weeks. This study underscores the potential of microalgal-based dietary supplements as natural, sustainable, scalable, and health-promoting alternatives in the context of modern, fast-paced lifestyles, where individuals seek convenient and nutrient-dense, fast options to support overall well-being.

## Data Availability

All data produced in the present study are available upon reasonable request to the authors

## 7. Data Availability

Data described in the manuscript will be made available upon request.

## Acknowledgement

SK, SV and RL designed research; SK and SV conducted research; DC analyzed data; and DC, SK, SV and RL wrote the paper. SK had primary responsibility for final content. All authors read and approved the final manuscript. Chlorella tablets are manufactured and supplied by EID parry, Nutraceuticals. We express our sincere gratitude to *Saptagiri Hospitals* for their valuable support and cooperation in the successful completion of this work. We also thank Ms. Saumya Subramanian for her contribution in reviewing and editing the manuscript. We acknowledge the use of OpenAI ChatGPT *(version 5)* for minor language editing. However, we take full responsibility for the scientific accuracy and integrity of the manuscript.

