## supplementary file for "Assessing the efficacy of Chlorella vulgaris for Vitamin B12 Deficiency: A Randomized Controlled Trial"

eurofins

**Batch code:** EUINBA-00157578
**Report code:** AR-23-IR-057637-01
**Report date:** 15.06.2023

ilac-MRA TL-1097 IAS ACCREDITED Testing Laboratory

**PARRY NUTRACEUTICALS, DIVISION OF EID PARRY(INDIA) LTD - OONAIYUR**
**PANANGUDI POST,**
**PUDUKKOTTAI DT,**
**622505OONAIYUR.**
**Tamil Nadu, INDIA**

Scan to authenticate this report

**Mr Balakrishnan RM.**

### ANALYTICAL REPORT

| | | | |
|---|---|---|---|
| **Sample code:** | 258-2023-06002383 | **Report code:** | AR-23-IR-057637-01 |
| **Sample name:** | Organic Chlorella Tablets | | |
| **Client Details** | 2 | **Received on:** | 08.06.2023 |
| | | **Analysed between:** | 08.06.2023 - 15.06.2023 |
| **Sample reference** | Customer Provided Details<br>Batch Number : TCTOD2301 | | |
| **Quantity received:** | 66g | | |
| **Sample packing:** | Sealed Aluminium Pack | **Sampling:** | NOT SAMPLED BY EUROFINS |

#### VITAMINS

| | | | *Method* | *Result* | *LOQ* | *Unit* |
|---|---|---|---|---|---|---|
| **IR199** | **IR** | Vitamin B12 (cyanocobalamin) | AOAC 2011.09 | 472.5 | 0.5 | µg/100 g |

The tests identified by the two letters code IR are performed by Eurofins Analytical Services India (Bangalore), INDIA.

**Ms Shalini Sharma**

**Manager - General Chemistry**

LOQ = Limit of Quantification

************** END OF REPORT ***************

The results may not be reproduced except in full, without a written approval of the laboratory. The results relate only to the sample analysed.
